# Inter-professional mentor teams (IP-MT) and systems analysis and improvement approach (SAIA) for elimination of mother-to-child transmission of HIV (eMTCT) at University affiliated teaching and referral hospitals in Kenya

**DOI:** 10.1101/2025.09.22.25336422

**Authors:** Ruth Nduati, George Wanje, Emmah Matheka, Ruth Emboyoga, Paul Nduati, Dalton Wamalwa

## Abstract

Elimination of mother-to child transmission (eMTCT) of HIV is a complex cascade of interventions longitudinally administered during pregnancy, delivery and after, until the baby is no longer at risk of infection. Systems Analysis and Improvement Approach (SAIA) an institutional implementation strategy that packages systems engineering tools have in clinical trial setting been shown to effectively optimize performance. This study evaluated feasibility of institutionalizing inter-professional mentoring teams (IP-MT) that utilize SAIA for strengthening delivery of eMTCT services at referral and teaching hospitals affiliated to the Faculties of Health sciences. Nurse midwives drawn from 15 referral and teaching hospitals affiliated to faculties of Health Sciences of 4 Universities in Kenya and who were trained on SAIA were tasked to mobilize fellow colleagues to build IP-MT to support eMTCT within their hospital following additional 2-day online SAIA training and follow-up mentorship as needed. In the current training 71 health workers of different cadres were trained. Retention of trained personnel was 37.5% and 92.9% for the first and second cohort respectively. Nine facilities established the IP-MT but only one hospital had participation of a consultant doctor. Eight hospitals were able to analyze their facility performance for the period preceding the training using the PMCT cascade analysis tool (P-CAT). Two-thirds of expected pregnant women booked into care. Four of the eight hospitals had consistent fidelity across all components of the eMTCT service. Two hospitals failed to provide anti-retroviral drugs to 50% of eligible women. Timely early infant diagnosis occurred in 271 (92.5%) of the 293 identified HIV exposed infants. Teaching and referral hospitals affiliated to University health professions schools offer opportunity for interprofessional collaboration and opportunity to improve the quality of eMTCT services. Collation of the P-CAT findings helped identify common program gaps and facility specific deficiency in delivery of eMTCT.

## 1 INTRODUCTION

The evidence from clinical trials and real-world settings is undisputable that elimination of mother-to-child transmission is feasible and indeed it is with this understanding that countries have committed to the Global Alliance for ending AIDS in children by 2030[1,2]. Towards this, in the Dar-es-Salam declaration of February 2023, twelve nations from East and Southern Africa including Kenya committed to achieve triple elimination of HIV, syphilis and hepatitis B virus infection by 2030 [1,2]. Prevention of mother to child transmission of HIV (PMTCT) is, a complex longitudinally administered intervention during pregnancy, delivery and after until the baby is no longer at risk of infection. Failure to deliver different components of PMTCT care cascade undermine the 2030 goal of eliminating pediatric HIV and AIDS. Kenya has a national prevalence rate of HIV at 4.0%, and an estimated 1.45 million people living with HIV [3]. The mother-to-child transmission rate (MTCT) in 2022 was 8.9% well above the elimination target of < 5% transmission rate. [2,4] In addition, up to 54.7% of HIV-infected children were born to women who had either discontinued or failed to initiate their ARV therapy [4].

Strategies shown to augment delivery and scale-up of eMTCT services include designating an interprofessional team to provide mentorship and support for; delivery of the prevention of mother to child transmission of HIV integrated with maternal-newborn and child health services (eMTCT/MNCH) [5], standardization of the care package, and optimization of service delivery using Systems Analysis and Improvement Approach (SAIA) to improve the quality of care. [6.7]. Systems Analysis and Improvement Approach (SAIA) is an evidence-based institutional quality improvement strategy which packages systems engineering tools to optimize care cascade performance of the eMTCT of HIV service [7]. SAIA is flexible to context and supports frontline staff gain a comprehensive view of their complex delivery systems, identify and prioritize areas to improve, and iteratively test modifications to increase system outputs and patient outcomes [8]. In clinical trial setting SAIA leveraged routinely collected data to inform decision-making in an authentic service delivery setting, resulted in a remarkable 3.3-fold increase in ART uptake for HIV-infected pregnant women and over a 17-fold increase in early infant diagnosis for infants exposed to HIV [7]. This adaptable and scalable model has been successfully adapted for several communicable disease conditions including pediatric [10,11] and adolescent HIV [12], HIV testing in family planning clinics (Kenya) [13], as well as non-communicable disease areas such as hypertension (Mozambique), cervical cancer screening (Kenya), mental health (Mozambique) [14], and opioid overdose reversal (US) [15]. SAIA like all other continuous quality improvement tools needs to be part of the institutions everyday function.

In Kenya University faculties of health Sciences partner with public hospitals for purposes of training. The role of University faculty of health sciences staff is guided by the institutions’ agreements with the hospitals which in our region are owned by different arms of government. Health facilities are organized into six categories; the first 3 levels are classified as primary care and the others are level 4, 5 and 6 as hospitals as capability increases. The first-generation health professions schools in Kenya are partnered with level 6 facilities, also known as National referral hospitals. There has been a gradual expansion of higher education, including establishment of a new generation of health professions schools that rely on less resourced regional hospitals (level 4 and 5) as training sites. In this context, the faculty role has been largely limited to patient care, teaching and research with minimal involvement in the continuous quality improvement activities of the hospitals. In the context of achieving the goals of eMTCT, the teaching and referral hospitals should be centers of excellence for service delivery and demonstration of good evidence-based practice. These teaching and referral facilities have large volumes of women seeking pregnancy and delivery services and failure to achieve targets significantly contributes to delays in achieving the set public health goals.

The University of Nairobi has a long-standing initiative to promote medical education at the teaching and referral hospitals where the health professions training takes place and as the oldest University health professions training institution, has reached out to the newer public training institutions. The HEPI program [NIH-FIC Grant No. 5R25TW011212-05] in Kenya which was designed to build capacity of faculty and health workers in hospitals affiliated to 4 partnering public universities faculty of health Sciences, in 2022, supported the training of nurse midwives who were direct service providers of eMTCT from 16 affiliated hospitals (two level 6, eight level 5 and five level 4 hospitals) on SAIA. The training provided a unique networking opportunity by bringing together teams of direct eMTCT service providers at these teaching and referral hospitals with the head of the National AIDS Control program (NASCOP). Discussions during the training revealed that there was great variability in the organization of the services, weak local ownership of the eMTCT and pediatric HIV services, over-reliance on non-governmental partner support and perception that HIV services are peripheral to the rest of the hospital. There were challenges related to staffing, knowledge gaps, weak linkages, and lack of commodity security. Institutional memory of effective service delivery strategy was tenuous contributed to by high staff turn-over and specifically among the nurse midwives who are core providers for maternal and child health. Establishing facility level inter-professional health teams that includes the affiliated medical school faculty was proposed as a strategy to promote these facilities to be centers of excellence in eMTCT and suited for their role in medical education.

We hypothesized that establishing an interprofessional eMTCT mentorship team and institutionalizing eMTCT-SAIA through continued medical education within each institution will provide a platform for ongoing improvement, program fidelity, and sustainability in the face of staff redeployment and diminishing resources. Our objectives were to evaluate feasibility of institutionalizing inter-professional mentoring teams (IP-MT) that utilize SAIA for strengthening delivery of eMTCT services at referral and teaching hospitals affiliated to the Faculties of Health sciences in 4 Universities in Kenya in close collaboration with the Ministry of health (MOH), County Health Departments and partner institutions supported by a grant, AFREhealth grant 1R25TW011217-05.

## 2 MATERIALS & METHODS

### Ethics Statement

The study aimed to improve prevention of mother-to-child HIV transmission through the institutionalization of SAIA and inter-professional mentorship practices with the expectation it would result in better health outcomes for mothers with HIV and their children.

*Autonomy* – Permission was sort from each of the hospitals to train the health-workers on SAIA in the context of delivering prevention of MTCT of HIV and to utilize the grouped service delivery data to monitor this quality improvement activity. Health-workers were invited to participate in the study. The study was granted a waiver of individual consent.

*Beneficence* – The inter-professional mentorship team approach and SAIA program aims to improve service delivery for prevention of mother-to-child HIV transmission at these teaching hospitals. The anticipated immediate benefits are that the health providers and their trainees would have improved knowledge of continuous quality improvement of the eMTCT service using the SAIA methods, be updated on the current national guidelines, and inter-professional mentorship and working relationships strengthened. There would be improved service delivery (efficient in-facility service delivery, better support for retention and compliance) to their clients and increased sustainability of eMTCT programs.

*Justice* – All health-workers delivering eMTCT services were welcome to participate in the training and follow-up CQI activity.

*Risks and benefits of the study -* Being a capacity building project, no risks were anticipated.

*Potential adverse events and proposed interventions -* The consistent challenge with HIV research is the risk of involuntary disclosure of HIV status. This study activities were focused on training health workers to conduct SAIA a quality improvement approach to the delivery of eMTCT services. The data was the summaries from the Kenya health Information system and the hospital’s own records and not individual patient data. Nevertheless there is a lingering risk that an innovation in service delivery may increase risk. *Minimization of Risk*: To mitigate against this included in the training of the health workers a review of the eMTCT guidelines, human subject protection training and certification (CITI) and encouragement to ensure high standards of patient care, and dignified health service provision. Researchers did not have direct contact with patients. Where we identified gaps in service delivery and linkage during review of the collated data, there was feedback to the team leads and brainstorming support on how to respond within the context of existing guidelines. *Unknown Conditions*: There is strong evidence that co-morbid conditions among care givers of HIV exposed and infected children impedes ART adherence and retention in care. During training on SAIA we highlighted the importance of addressing the co-morbid conditions as per the Kenya ART guidelines. We did not anticipate other problems.

*Treatment*: The study does not involve direct patient contact and therefore we did not anticipate to have adverse events.

*Financial Responsibilities*: At the end of the study period health workers were reimbursed at a uniform rate the cost of data bundles.

*Confidentiality of research data* The study engages with consolidated data and there are no individual patient identifiers. The data for the period we analyzed is stored in study computer with access limited to authorized personnel.

*Consent /assent forms and waiver* - In this study we requested and were granted a waiver of consent.

### Ethical approval processes

Ethical approval was given by the UON-KNH ERC P41/01/2024, and NACOSTI research permit number P/24/35987 as per the Kenya government regulations.

*Study Design* - This is a hybrid type 3 implementation study to explore feasibility and acceptability of SAIA to facilitate optimization of eMTCT at Maternal newborn Child Health (MNCH) services of [16,17].

*Study setting **-*** Referral and teaching hospitals affiliated to the Faculties of Health sciences in 4 Universities in Kenya (University of Nairobi, Kenyatta University, Jomo Kenyatta University of Agriculture and Technology and Maseno University).

*Inclusion and exclusion criteria* – Eligible facilities were included if the in-charge accepted to participate. Within the institution health workers who directly work on maternal child health services where prevention on mother-to-child services are integrated in the ANC, maternity and in the well child/well woman clinic as managers or as direct service providers were included. Those eligible for leadership roles in the facility by virtue of their training were prioritized for the training. Facilities and health workers who were reluctant to participate were excluded from the study.

*Recruitment Process:* The 32 PMCT providers previously trained on SAIA were tasked to mobilize fellow colleagues towards building inter-professional mentorship teams. Other than a phone call followed up with email communication, no additional training was provided for this task. In each facility leadership and staff who provide PMCT services were invited to volunteer to establish SAIA-inter-professional mentorship teams (SAIA-IPMT). Guidance was provided that membership would include all cadres involved in providing eMTCT services.

*Training Approach* The nascent (SAIA-IPMT) teams were provided with an online course on SAIA contextualized to delivery of eMTCT. The understanding was that the team would then be supported to utilize this approach within their institution to improve performance and resilience of the eMTCT services. The training was organized into 6 standardized modules; Introduction into Applied Systems Engineering Approaches for HIV Service Improvement, Introduction to Systems analysis and improvement, SAIA methods 1 Cascade Analysis, SAIA methods 2 - Value Stream mapping; PDSA cycle and Data collection to support SAIA. These modules were facilitated online by GW, RW and EM over a period of 2days and follow-up mentorship provided by the same team.

*Systems analysis intervention action (SAIA)* - SAIA is a five-step process that draws from systems engineering in the Toyota Production Systems and research in low- and middle-income countries (LMICs) to improve quality of care [5, 6]. Step 1 involves the use of an Excel-based tool to quantify drop-offs or individuals who did not progress in each step of a process, allowing users to identify and improve one step in the cascade while holding the other steps constant as shown in figure 2. [7, 8]. Figure 2 shows the output of the PMCT cascade analysis tool (P-CAT), which is formulated into a google store App.

**Figure 1.**
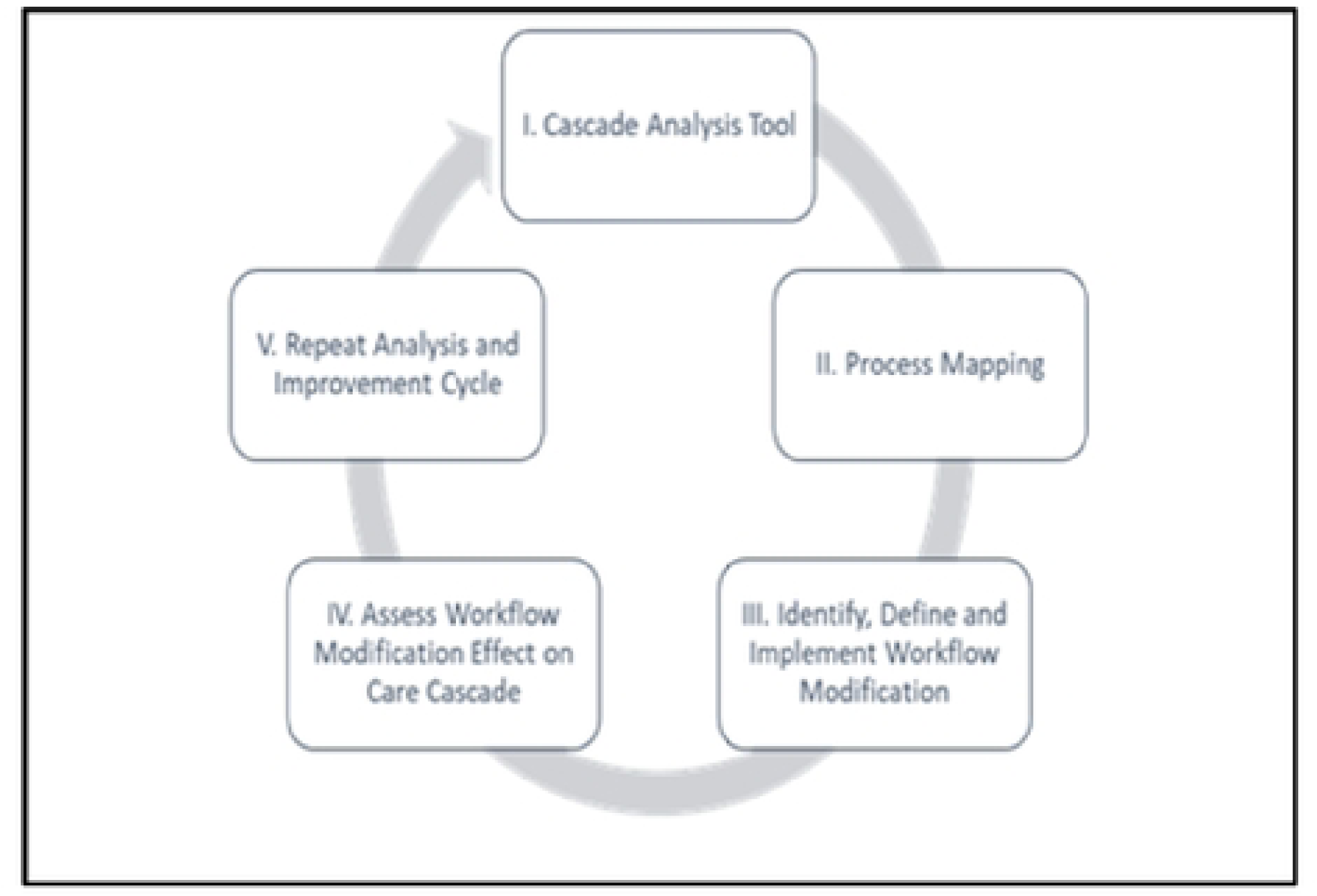
This is the Fig 1 Title. SAIA Core components

**Figure 2.**
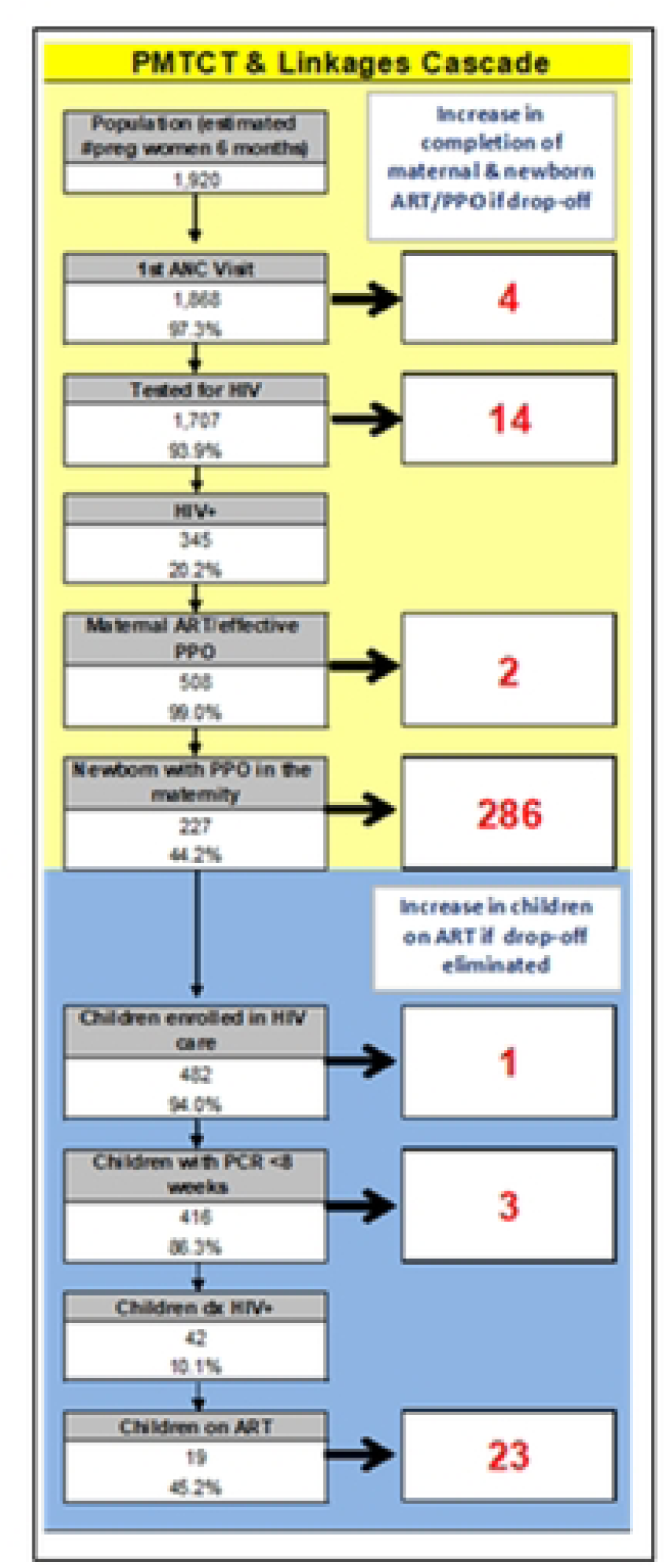
This is the Fig 2 Title. Example of the PMCT cascade tool (PCAT)

Step 2 involves process flow mapping with clinic staff to identify modifiable bottlenecks redundancies in delivery of the key areas for intervention. In step 3, a workflow modification is developed and implemented to address the bottleneck identified in step 2, constituting a continuous quality improvement (CQI) step. Step 4 assesses the impact of the modification and recalculates the cascade analysis in step 1 (another CQI step). Step 5 repeats the cycle for CQI. The teams were then tasked to develop a care cascade and flow chart for their facility. The SAIA care cascade excel spread sheet was provided to the teams to fill in their data. Online coaching and site visits were provided for facilities that struggled with achieving this task. The teams were expected to develop their strategies for optimizing delivery of eMTCT services within their institution.

*Assessment of outcomes*: The RE-AIM model [18] was used to evaluate the programs. Reach was measured by number of facilities reached, effectiveness was measured by the number of staffs that were trained, Adoption was manifest by whether facilities were able to form the IP-MT. Implementation was manifest by ability and willingness to construct the P-CAT and sharing the same with the researchers while maintenance is partly measured by proportion of trained staff retained and the provision of eMTCT services. The outcome indicators for the program included the achievement of the set targets at each step in the provision of services from pregnancy through to point of first evaluation of infant HIV infection status. The data sources were (i) training records (ii) minutes of the SAIA iterative meetings at the participating sites aimed at collating data and reviewing impact of the quality improvement interventions the site had made and (iii) the care cascade based on the routinely collected daily activity register in the ANC, delivery and postnatal HIV exposed child registers. The research team has aggregated the data from the different PCATs for this analysis.

### Data analysis

A simple count and stratification by cadre is used to demonstrate the formation IPMT. Proportions are used to describe staff retention and success in adoption of the PCAT tool. A cascade analysis is used to present the adoption of this process by the institutions. The first step of the PMCT care cascade is booking into antenatal clinic. In order to make this cascade the facility needed estimates of their catchment population and expected number of pregnant women for the year. The facilities were requested to obtain these estimates from the local office of statistics (an arm of the ministry of Planning) which are based on census data and official projections of population. Uptake of HIV testing was computed as number of women who accepted HIV testing as a percentage of those who had unknown status at point of booking into ANC. Facility HIV prevalence was computed as total positive women (known positives and newly diagnosed) as a proportion of the total number of women booked into ANC during the period of observation. Prevalence of HIV among women diagnosed during the index pregnancy was computed as women newly diagnosed with HIV as proportion of women who were not known positives and therefore offered testing as part of the ANC care package. A 95% confidence interval was computed for the HIV prevalence estimates. The first infant testing should be by 8 weeks of life. HEI access to diagnosis and timeliness of the testing was analyzed in two different ways, the first a longitudinal analysis using the number of HIV infected women as denominator for determining coverage, and the other analyzing infant data as a separate cohort. One facility which had large inconsistences in the reported PCAT data and therefore the longitudinal analysis was done with and without this facilities data on the premise that the inconsistences are indicative of the gaps in the program delivery, and risk of failing to meet targets. In the discussion, the estimates from this study are used to project the number of women and infants who suffered a missed opportunity for eMTCt services.

## 3 Results

The 15 teaching and referral hospitals affiliated to the 4 faculties of health Sciences are distributed across 10 counties in Kenya as shown in figure 3 and these included two level 6 national referral and teaching hospitals, eight regional referral and teaching hospitals and five County level hospitals. All 15 hospitals participated in the first round of SAI training in 2022 and 10 participated in the second training in 2024. The first SAIA training reached 32 staffs all of them nurse midwives. Five facilities that did not participate in the second round of SAIA training were regional referral facilities. The 10 facilities that participated in the second round of SAIA training, 4 had the 2 previously trained staff still engaged in delivery of eMTCT services, while the other 6 had only one previously trained staff participating. In the second round of training 71 staff were trained but before the completion of the project 9 had already left from transfers and retirement. Staff retention was 12/32 (37.5%) over 2 years from the first training and 92.9% over one year following second training.

**Fig. 3.**
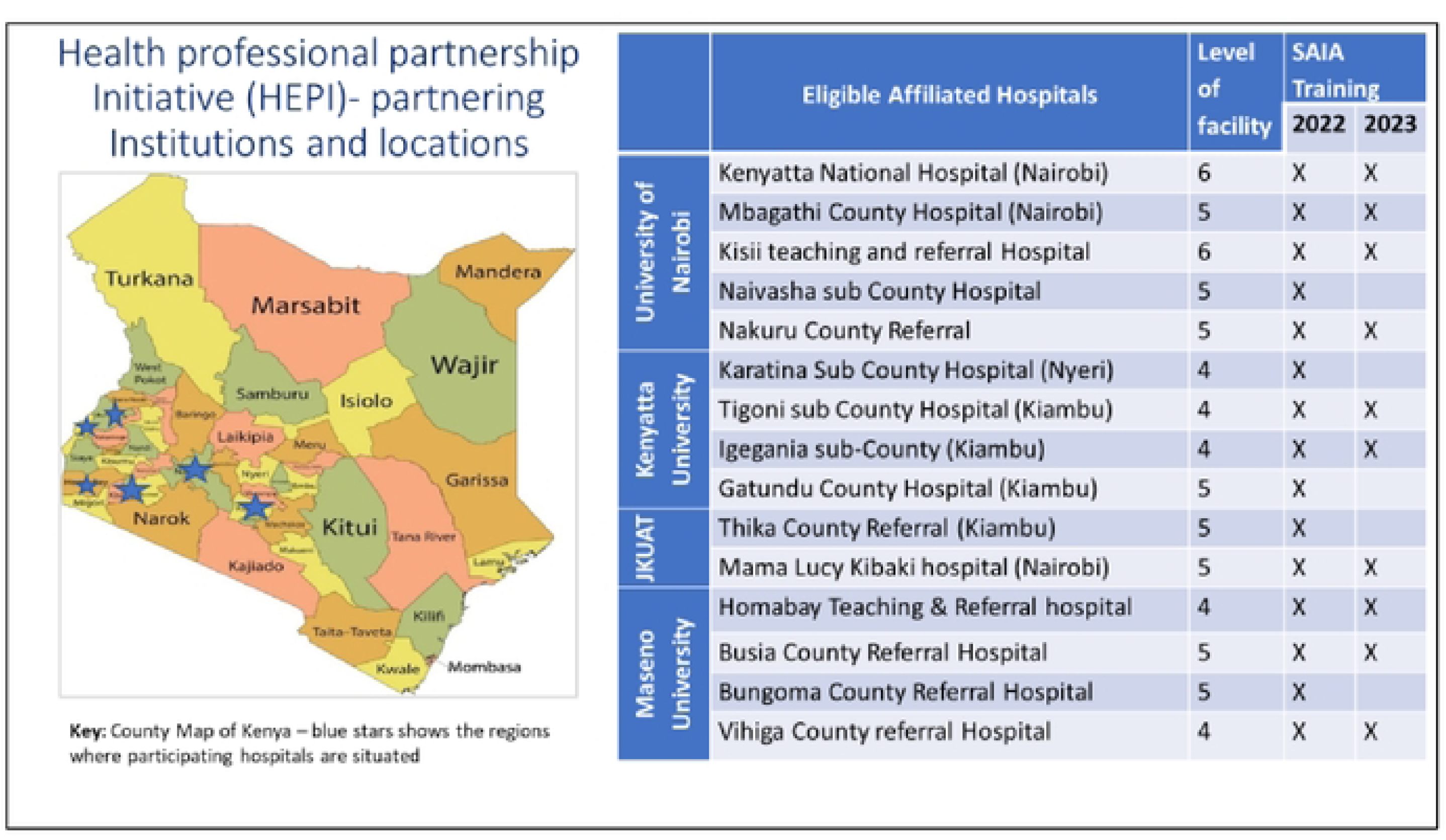
This is the Fig 3 Title. Distribution of the study sites

Nine of the 10 teams formed a SAIA inter-professional team. Eight teams were then able to construct the P-CAT using the routine facility levels records. Figure 4 below shows the cascade of adoption of SAIA in these 16 facilities.

**Fig 4.**
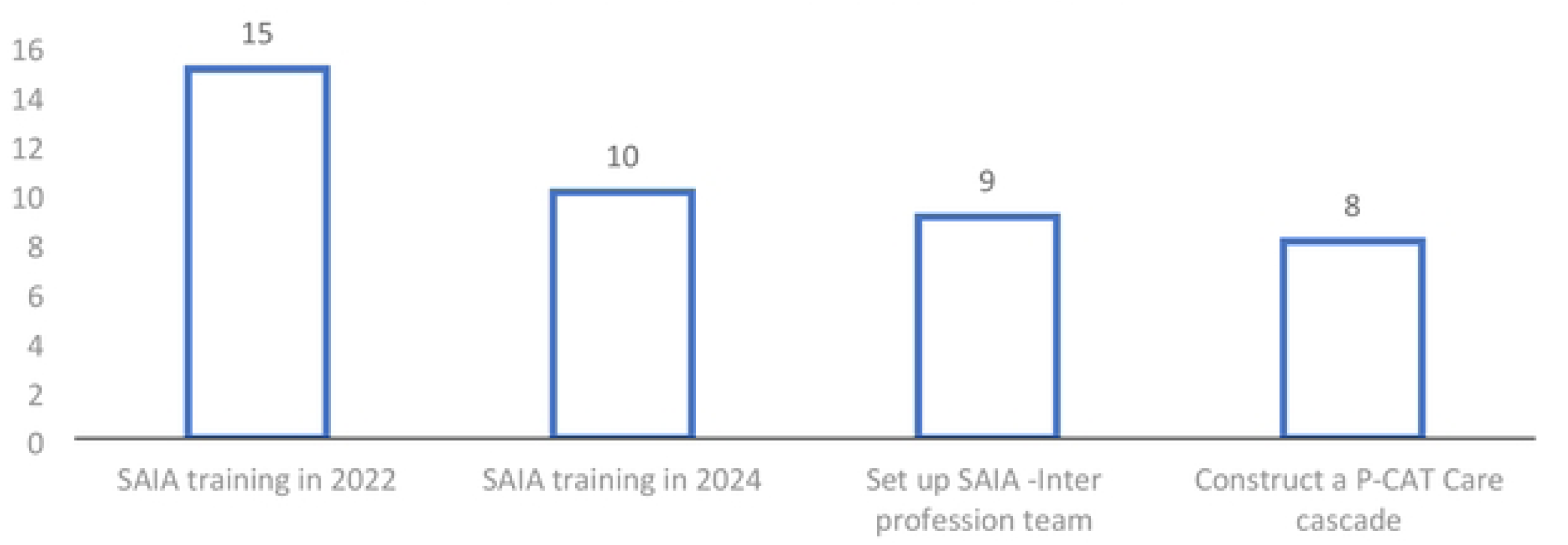
This is the Fig 4 Title. Uptake of SAIA by the University affiliated hospitals

### Structure of the SAIA Teams

Eight of the facilities shared the structure of their SAIA teams. There was an average number of 9 members per team (range 6-15). The majority in each team were the nurse midwives who in addition to direct service provision played various leadership roles in providing eMTCT services as shown in figure 5. Seven teams included a medical doctor, but only one included a consultant level medical doctor, a paediatrician. All but one hospital had a pharmacist, laboratory technologist and a nutritionist as members of the team. Six of the 8 facilities incorporated the medical records officer. One level 6 facility with a high case load incorporated the HIV testing staff, and peer mentor as members of the IP-MT.

**Fig 5.**
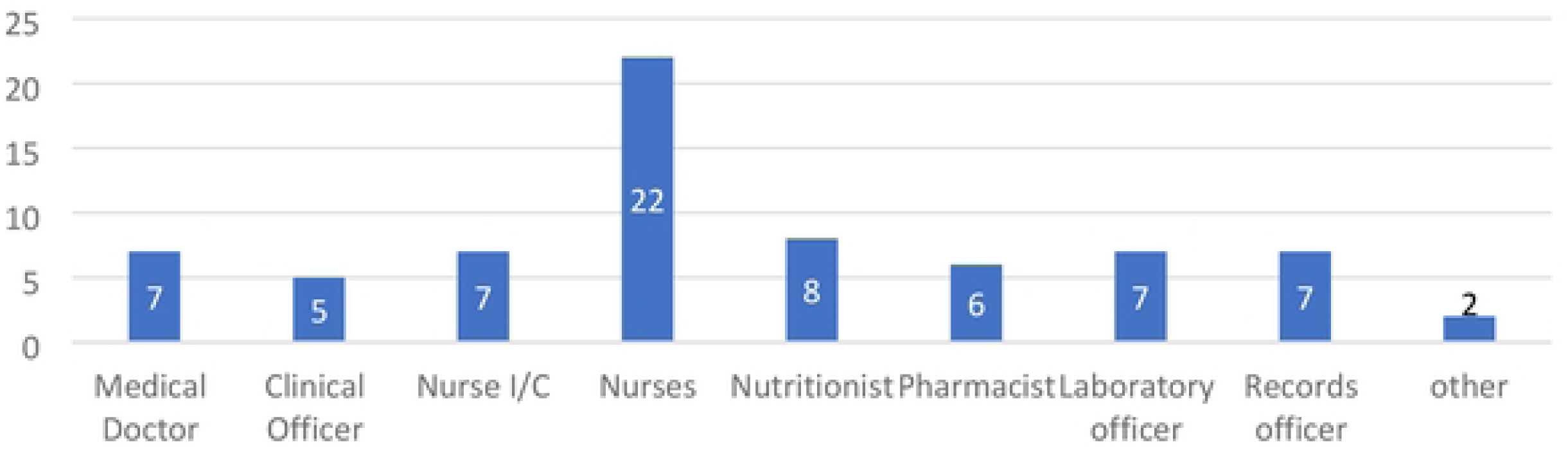
This is the Fig 5 Title. Cadres of health-workers in SAIA inter-professional teams

**Fig 6.**
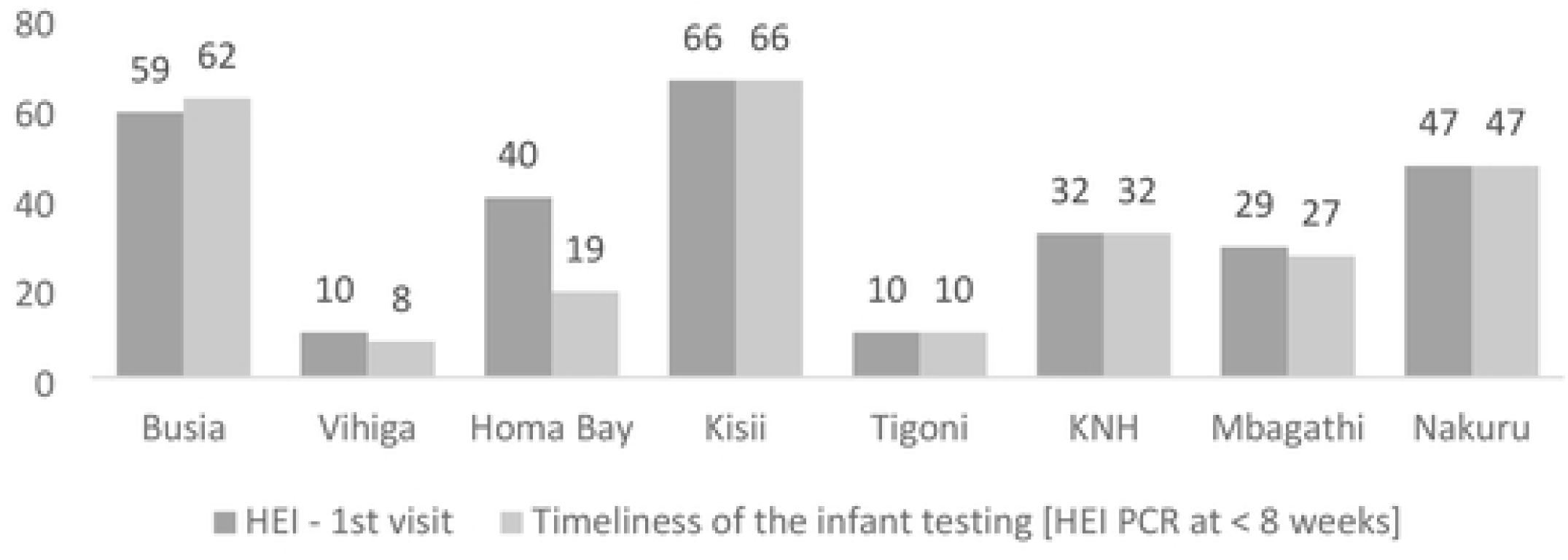
This is the Fig 6 Title. Comparison of the number and timeliness of HIV exposed infant visits

### Construction of the PMCT Care cascade using the Excel P-CAT Template

Seven facilities reported on 4 months of observation and one on 6 months as shown in Table 1. Facility 8 had challenges in constructing the first step of the care cascade. Data from this facility is retained in the analysis to illustrate challenges faced in achieving the eMTCT targets.

**Table 1:** Step 1 of Prevention of mother to child transmission of HIV (PMCT) care cascade.

| Facility | Catchment population | Estimate used to compute number of pregnant women | Duration of observation Jan-April 2024 in months | Target population for period of observation | Step 1 - seeking ANC care |  |  |  |
| --- | --- | --- | --- | --- | --- | --- | --- | --- |
|  |  |  |  |  | Seen as 1st ANC | New ANC client/month | % of target population seen at first ANC | Estimated Absolute number missed |
| Busia | 79961 | 3.1% | 4 | 826 | 621 | 155 | 75% | 222 |
| Vihiga | 35462 | 5% | 4 | 591 | 212 | 53 | 36% | 379 |
| Homa Bay | 33882 | 5% | 4 | 528 | 442 | 111 | 84% | 86 |
| Kisii | 60336 | 5% | 6 | 1508 | 1193 | 199 | 79% | 315 |
| Tigoni | 9295 | 5% | 4 | 156 | 270 | 68 | 174% | 0 |
| KNH | 436 | 5% | 4 | 436 | 436 | 109 | 100% | 0 |
| Mbagathi | 115,685 | 5% | 4 | 1928 | 331 | 83 | 17% | 1597 |
| Nakuru | 71,186 | 5% | 4 | 1186 | 980 | 245 | 83% | 206 |
| **Total** | 406243 |  |  | 7159 | 4485 | 1121 | 63% | 2805 |

*Step 1 of prevention of mother-to-child transmission of HIV is access to the antenatal services*: The average number of newly booked ANC clients ranged from 53-200 per month. In the first re-iteration, six facilities failed to reach the projected target of pregnant women. Tigoni a facility with a large catchment of migrant farm workers had a 74% increase over the expected new antenatal clients. The proportion of women reached and the absolute numbers of the expected women who did not book ANC are shown on 8^th^ & 9^th^ column of table 1. Overall 2805 (37%) of the expected 4485 women did not book into ANC.

*Step 2 Uptake of antenatal HIV testing*: Seven facilities had sufficient to data to enable evaluation of this step. Five facilities reached all the women with unknown HIV status with HIV testing. Busia a border town facility had 108% uptake while Tigoni a facility with a large catchment of migrant farm workers achieved 95% testing rates. Excluding Nakuru that had insufficient data on step 2 there were 185 women with previously known HIV positive status and 52 who were newly diagnosed making a total of 237 cases. The overall HIV prevalence was 237 [6.8% (95% CI 5.97,7.64)] for the 3505 women who booked into the 7 facilities as shown on table 2. Excluding the known positive women, 52 (1.54 (95% CI 1.18,2.03%) of 3355 women offered HIV testing in the ANC were found to be HIV infected. The facility level HIV prevalence among newly tested clients and the 95% Confidence interval is displayed in table 2. In 4 facilities, the HIV prevalence in the newly diagnosed women was significantly lower than the overall facility HIV prevalence as demonstrated by discreet confidence intervals.

**Table 2:** Step 2 of the PMCT care cascade on access and uptake of HIV testing.

| Facility | Number of KPs<sup>1</sup> | Step 2: HIV testing |  | Number new identified Positive | Facility level HIV Prevalence among newly diagnosed women % (94% CI) | Overall HIV prevalence among women booking ANC % (95% CI) | Total HIV positive women reached in the ANC clinics | Step 3 | Step 4 |
| --- | --- | --- | --- | --- | --- | --- | --- | --- | --- |
|  |  | STEP 2: ANC tested | % of eligible population at ANC |  |  |  |  | HIV positive women who receive ARV before or at and after delivery | Number of women issued infant prophylaxis |
| Busia | 47 | 621 | 108% | 4 | **0.64% (0.25,1.64)** | 9.0% (7.01,11.53) | 56 | 56 (100%) | 56 (100%) |
| Vihiga | 17 | 195 | 100% | 18 | 9.23% (5.91,14.12) | 16.5% (11.12,22.0) | 35 | 18 (51.4%) | 18 (51.4%) |
| HomaBay | 44 | 398 | 100% | 6 | **1.51% (0.69,3.25)** | 11.31% (8.69,14.6) | 50 | 50 (100%) | 50 (100%) |
| Kisii | 33 | 1160 | 100% | 13 | **1.12% (0.66,1.91)** | 3.85% (2.90,5.10) | 46 | 46 (100%) | 46 (100%) |
| Tigoni | 7 | 251 | 95% | 7 | 2.79% (1.36,5.6) | 5.57% (3.21,8.96) | 14 | 7 (50%) | 7 (50%) |
| KNH | 30 | 406 | 100% | 2 | **0.49% (0.14,1.78)** | 7.3% (5.24,10.18) | 32 | 32 (100%) | 32 (100%) |
| Mbagathi | 7 | 324 | 100% | 2 | 0.62% (0.17,2.2) | 2.72% (1.44,5.09) | 9 | 9 (100%) | 9 (100%) |
| Nakuru | 143 | 9 |  | 9 | missing data |  | 152 | 380 | 9 |

<sup>1</sup>Known positives

*Step 3 and 4 Provision of anti-retroviral drugs to the HIV positive women and prophylactic doses for their infants:* The current policy is to initiate ART on the day of diagnosis for newly diagnosed women and at the same time issue infant ARV prophylaxis dose to enable delivery of the infant prophylaxis within the 72-hour window after delivery. A total of 218 (90%) of 242 HIV positive women (excluding Nakuru) were issued ARV’s for themselves and their infants. Five facilities issued ARV drugs to all women identified as HIV infected as shown in Table 3. In 2 facilities half of the women missed receiving ARV drugs for themselves and their infants, 18 [51.4% 95% CI 35.6,67)]of 35 women in Vihiga and 7 [50% (95% CI 26.8,73.2)] of 14 women in Tigoni as shown on table 2. Nakuru a regional referral facility reached 152 HIV positive women through their antenatal clinics (145 known positive and 9 newly diagnosed clients) but reported that they provided ARV’s to 380 mother-infant pairs. In all instances infant ARV prophylaxis was issued at the same time as the mother’s drugs.

**Table 3:**
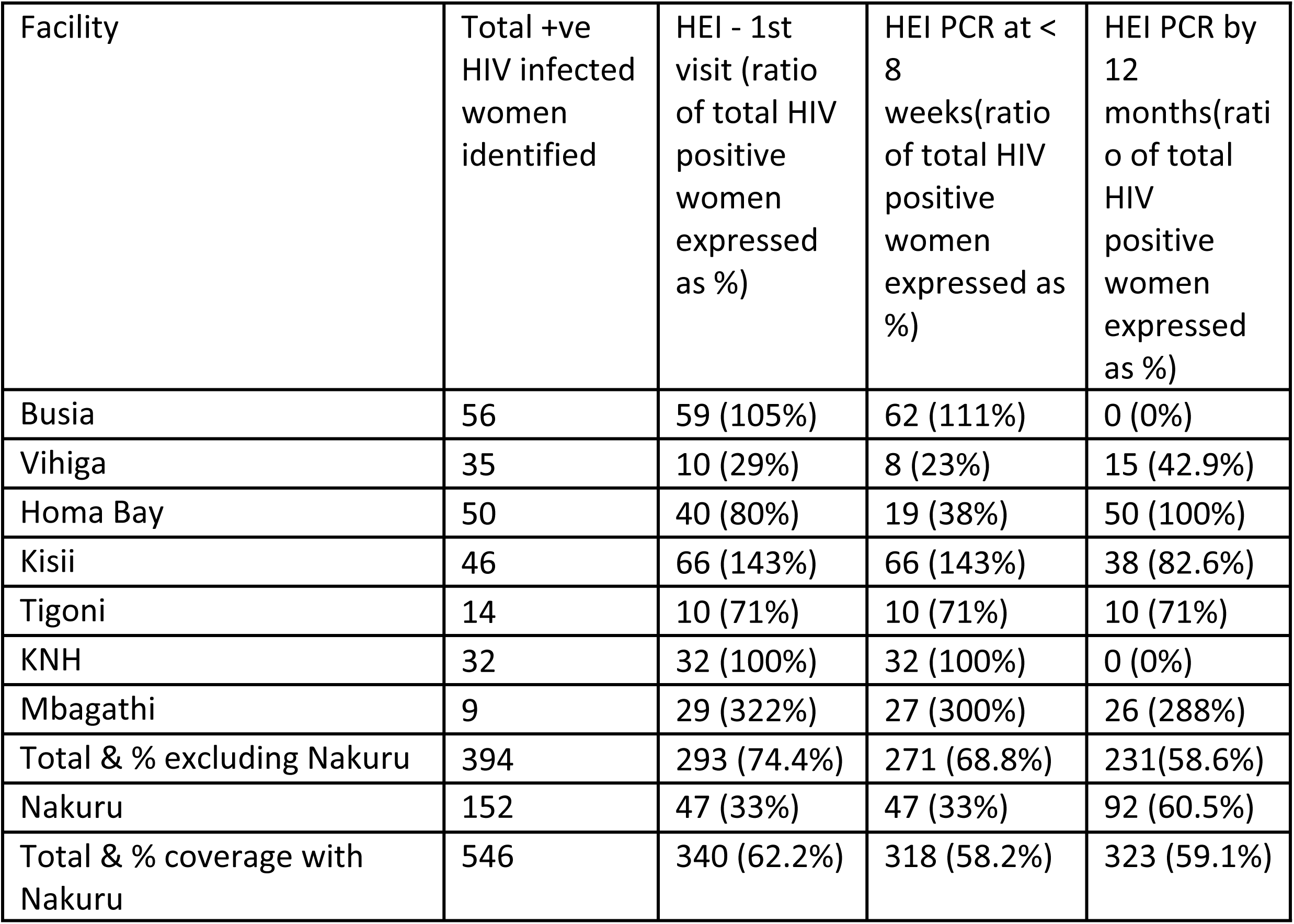
Access to early infant diagnosis.

**Table 4.**
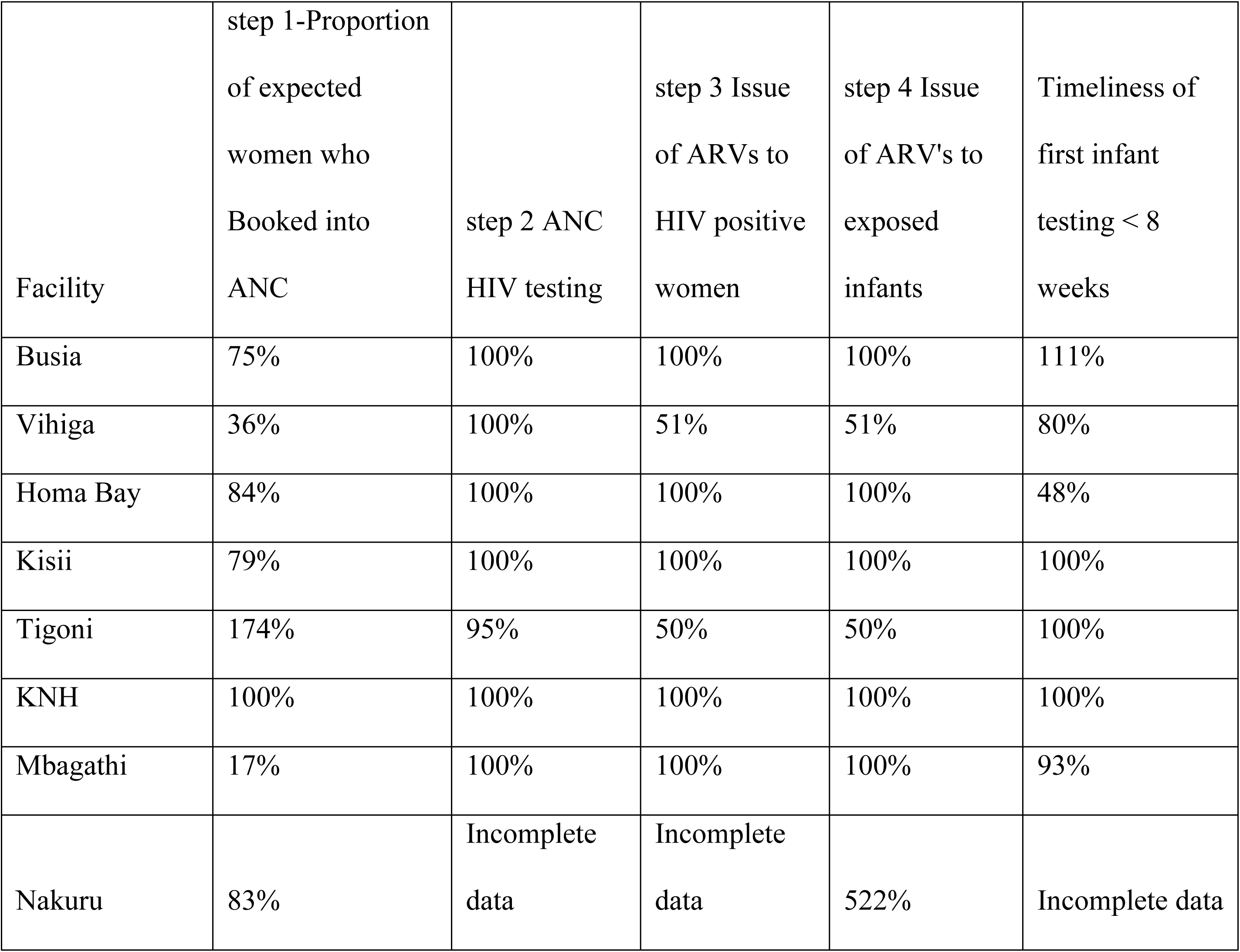
Proportion of women and infants receiving care at each step of the care cascade.

*Early Infant diagnosis (EID)***: -** As expected the number of HEI at first visit do not match the numbers of HIV infected women in the facility. Excluding Nakuru data a total of 293 (74.4%) infants were seen for a first HEI diagnosis among them 271 (68.8%) who had a timely test at < 8 weeks of life. When data from Nakuru is included the estimated proportion of HEI who accessed testing and the timeliness dropped to 62.2% and 58.2% respectively. One may argue that since the data for this analysis is based on daily activity registers the HEI could be considered as an independent cohort.

In this analysis where infant testing is treated as a separate cohort from the mother’s care cascade. Overall 92% of the infant had a timely early infant diagnosis. (EID). In three facilities Vihiga and Homa Bay and Mbagathi, 80%, 48% and 93% of the HEI had a timely diagnosis at < 8 weeks of life respectively, lower than the program target of 95%. The other 4 facilities had 100% timeliness of HEI testing. None of the

HEI returned to the National hospital for the 12 months visit in compliance with current policies while County Referral hospitals had an influx of HEI infants.

*Timely initiation of anti-retroviral treatment* – In the study time period 3 (1%) of the 293 HIV exposed infants were identified as HIV infected but only one is recorded to have been initiated on treatment as required.

*Longitudinal assessment of performance per facility. -* Figure 5 shows the performance of the facilities for the different eMTCT services. During this period of time the weakest point in the delivery of elimination of MTCT of HIV was access to antenatal care where 6 of the 8 facilities failed to reach the targeted population. Two facilities had extremely low reach of anticipated ANC population 17% and 36% respectively. The only step performed perfectly in all facilities was the antenatal HIV testing that achieved the target of ≥ 95%. Two facilities failed to provide infant prophylaxis to half of the women at the time maternal drugs were issued. Two other facilities had a low access and timeliness of early infant diagnosis.

## 4 Discussion

We show that inter-university health professions schools forums have the potential to build capacity and networking opportunities for affiliated teaching hospitals by providing for an inter-professional training environment that address relevant public health problems. The affiliated teaching hospitals were willing to try out SAIA as a method of optimizing delivery of eMTCT services. The commitment to the process is demonstrated by release of staff to train and staffs willingness to attended the online SAIA training investing personal time and resources. In this study 9 teams succeeded in establishing an inter-professional SAIA team with membership of all the different players; doctors, nurses, laboratory technicians, pharmacists, nutritionists, health records officers and community promoters. SAIA is a monitoring and evaluation tool at point of service and at the same time a management tool which can be used to increase institutional ownership of the program. The consultants in the related fields of obstetrics and gynaecology and paediatrics and child health are knowledgeable about HIV medicine however only one SAIA team had a consultant level staff a paediatrician. In the environment of shortage human resource for health few medical specialists are deployed to serve in-patients and specialty clinics. Highly structured out-patients service such as antenatal care, TB, and HIV clinics and where care package is highly standardized are almost entirely managed by mid-level cadres, nurses and cadres. Medical specialists form part of the senior management team in a hospital. Their absence from the IP-MT gives the impression of lack of ownership of the eMTCT services. There may be real or perceived barriers to specialist involvement, some of it probably influenced by the structure of specialist which is almost entirely based on clinical services with minimal exposure to quality assurance processes of the teaching hospital.

This study is also illustrative of the challenges and obstacles to reaching the 2030 goal of eliminating new infant infections. The first problem was that one in three pregnant woman did not show up at the door. This is inconsistent with the Kenya Demographic Survey 2022 that shows that the regions where this study was done, 98% of pregnant women have at least one ANC visit with a skilled provider, and more than 85% have a skilled delivery which in nearly all instances is a health facility. [12] The study was conducted at the tail-end of a nation-wide rapid results initiative (RRI) for the country’s eMTCT program and whose objective was to optimize care and clean-up the monitoring data and therefore we have confidence in the reported data. There are several possible reasons for our findings. The observed deficit in antenatal bookings could be an indication that assumptions made in estimating the facility catchment of pregnant women are not cognizant of the decline in fertility rates. [12] Since 1989 Kenya’s total fertility rate has declined from 6.7 to 3.4 and in urban areas where these teaching hospitals are, TFR declined from 4.5 children to 2.8 children in the same time period. Kenya has rolled out Universal health coverage and in-built are economic benefits for utilizing lower level facilities (dispensary and health centers) and which may explain the deficit of antenatal mothers at the study hospitals which are level 4, 5 and 6 referral and teaching facilities.

Step 2 access to HIV testing was very well done and this excellent performance maybe a true indication of the service uptake, but also maybe a function of when a person is entered onto the register. If completion of the antenatal profile is a pre-requisite to be registered then women who decline to test may fail to be documented and indeed maybe may partly explain the disconnect between estimated number of antenatal clients and the ones who showed up, an observation made on a few process maps presented during the training.

If we assume the deficit in antenatal mothers is real and that their risk factors for HIV infection are similar to the women who showed up, applying the documented HIV prevalence to this population, there are 190 HIV infected women who did not book into ANC. In a worst case scenario where all of HIV positive women are not taking their ARV’s and assuming MTCT rate of 36.7% which has been previously documented in ARV naïve breastfeeding mothers, there would be approximately 68 infant HIV infections (20]. If on the other hand we assume the ratio of KP’s to new diagnosis found in this study applies to the women who did not show up in the antenatal clinic, 42 (22%) of the 190 babies would have been born to newly diagnosed women and we estimate they would contribute 15 infant infections. The true number of undiagnosed new infant infections lies somewhere between these estimates, and greatly influenced by the level of viral suppression among the KPs among the women who fail to book into antenatal clinic. With the same assumptions of HIV prevalence, we estimate that a further 3 infant HIV infections from the women who were diagnosed with HIV but failed to be started on ART.

Step 3 which is issue of ARV’s to the mother for herself and step 4 for issue of infant prophylaxis matched perfectly in the data for 7 of the 8 facilities showing that the providers were adhering to recommended best practice of issuing infant prophylaxis at the same time as mother’s ART. Overall 213 (92%) of 237 HIV positive were issued with ARV’s for themselves and their infants short of the 95% target. Two facilities, a quarter of the 8 facilities in this study, one in every two HIV infected women and their babies were documented to have not received their anti-retroviral drugs. National data shows 50% of new infant infections are distributed evenly between newly diagnosed women who fail to receive their ART and KP’s who have stopped taking their drugs. [3,4]

Showing up for early infant diagnosis (EID) in a timely manner is another key eMTCT step. In this time period 318 had a first HEI testing among them 271 (85.2%) accessed timely testing by 8 weeks of life as per the Kenya guidelines. Three (1%) of the 394 HIV exposed infants were identified as HIV infected but only one is recorded to have been initiated on treatment as required. These referral and teaching facilities experienced an influx of patients beyond their estimated targets for EID.

The strengths of this study is that it represents both urban and rural populations as well as teaching hospitals for Faculties of Health Sciences for 4 different Universities. These health facilities offer a diverse experience in terms of size, staffing norms, HIV prevalence and population served. Five are in regions with HIV prevalence above the national average, while other serve large populations of urban poor and others rural populations. This study shows how more granular data at facility level identifies the exiting gaps in service delivery Involvement of the teaching hospitals provides opportunities for pre-service and in-service training of the wider health work force and thus widening acceptance of the strategy and improvement of care. Success in eMTCT requires consistent fidelity to all the different components. Other than the national referral hospital, all other facilities face different level of challenge in providing the components of eMTCT that require longitudinal follow-up. These weaknesses are probably not unique to the eMTCT program and may reflect wider systems weaknesses in chronic disease follow-up. The stakes in HIV disease follow-up are high, in that a missed step often translates to a new infant infection or missed early diagnosis and timely treatment initiation. The majority of HIV positive women served in these facilities are women who have previously been diagnosed with HIV. The facilities need to have the capacity to provide sufficient adherence counseling to newly diagnosed women as well as support for the known positives. The antenatal clinics need to invest in treatment and adherence counseling to support this diverse population of women living with HIV through pregnancy and through the breastfeeding period.

A weakness of the PCAT is that one is not able to differentiate between infants of women accessing care at the facility versus those referred in for different components of the eMTCT program. The data from Nakuru is illustrative of this weakness. In this facility there were many previously diagnosed women (KP’s) and a big accumulation of HIV positive women at step 3. Including Nakuru data 394 HIV positive women were reached of whom 227 (57.6%) received the ARV’s. If one excluded the data from Nakuru, 90% of the identified positive women received their ARV’s. The truth lies somewhere between these two figures and it demonstrates the critical need for facilities with large numbers of pregnant women making sure they reach all women with services that they require along each step None of the facilities achieved the 95-95-95 percent targets for testing, treatment initiation.

A further limitation of the study is that the data is based on daily activity registers and not on longitudinal follow-up of the individual client. Such summary data may give a false sense of security for the program. The health sector is progressively moving towards individualized electronic medical records. In the future it will be possible to really have a patient level follow-up and which will facilitate identification of the vulnerable populations for additional support.

## 5 Conclusion

Prevention of mother-to child transmission of HIV (PMCT) PMTCT is, a complex longitudinally administered intervention during pregnancy, delivery and after until the baby is no longer at risk of infection. A multi-disciplinary team is needed to provide this service including clinicians, nurses, laboratory, pharmacy, supply chain and community members. These teaching facilities involved in this study were able to set up interprofessional teams. The teams succeeded in constructing a care cascade. The teams were also willing to share their data with the team. The P-CAT cascade shows in a snap-shot the strengths of the program as well as the opportunities for improvement. The greatest gain towards eMTCT of HIV will be in addressing local bottle necks in booking for antenatal care. Faculty at teaching and referral hospitals are untapped resource for quality improvement for the hospitals.

## 6 Recommendation

The inter-professional SAIA program should be adopted and institutionalized as part of the routine quality improvement tool that provided specific guidance on the priority steps in service delivery and the attainment of the goal of elimination of mother-to-child HIV transmission services and other conditions needing chronic follow-up. Greater effort should be made to involve the more senior doctors in a facility including the faculty from the health profession school’s faculty as a strategy for improving the implementation and ownership of SAIA and the eMTCT program and its targets. The current care cascade demonstrates low access of antenatal services. This is a direct threat to all the protection provided to pregnant women and their newborns when they access antenatal services. A priority action would be to pull together a community dialogue to find out what challenges the community is experiencing in accessing antenatal care.

## Data Availability

Data for this study is available on request from the communicating author. The same data is publicly available from the Kenya Ministry of health HIV data base

## Acknowledgement

We acknowledge the participation of the health workers and the health facilities included in this activity and the project coordinators in the department of Paediatrics and Child Health at the University of Nairobi.

## 7 Acknowledgements

We acknowledge the input of Dr Patrick Mburugu from Kenyatta University of Science and Technology in the conceptualization but was unavailable to contribute to the execution and write up of this article. Both contributed to the initial discussions regarding the proposal.

## Author contribution

Ruth Nduati, Conception and design of the study, analysis and interpretation of the data, drafting of the manuscript

George Wanje, Conception and design of the study, acquisition of the data, creation of software used in the analysis

Emmah Matheka, Conception and design of the study, interpretation of the data, review of the manuscript

Paul Nduati4 Analysis and interpretation of the data, review of the manuscript

Ruth Emboyoga, Conception and design of the study, acquisition of the data, review of the manuscript

Dalton Wamalwa Conception and design of the study, review of manuscript

## 9 SUPPORTING INFORMATION

**S1** Fig 1. **This is the S1 Fig 1 Title.** SAIA Core components

**S2 Fig 2. This is the S2 Fig 2 Title.** Example of the PMCT cascade tool (PCAT)

**S3 Fig. 3. This is S3 the Fig 3 Title.** Distribution of the study sites

**S4 Fig 4. This is the Fig 4 Title.** Uptake of SAIA by the University affiliated hospitals

**S5 Fig 5. This is the Fig 5 Title.** Cadres of health-workers in SAIA inter-professional teams

**S6 Fig 6. This is the Fig 6 Title.** Comparison of the number and timeliness of HIV exposed infant visits

**S1 Table. This is the S1 Table Title**: Step 1 of Prevention of mother to child transmission of HIV (PMCT) care cascade

**S2 Table This is the S2 Table Title**: Step 2 of the PMCT care cascade on access and uptake of HIV testing.

**S3 Table This is the S3 Table Title**: Access to early infant diagnosis

**S4 Table This is the S4 Table Title**: Proportion of women and infants receiving care at each step of the care cascade.

## Notes

### Competing Interest Statement

The authors have declared no competing interest.

### Funding Statement

The research work that is reported here was funded by AFREhealth grant 1R25TW011217-05 and the Health-Professional Education Partnership Initiative (HEPI) – Kenya NIH-FIC Grant No. 5R25TW011212-05. The report reflects the opinion of the authors and not of the funding agency or the sponsors of the health facilities.

